# Persisting Psychological Complications Following the Use of Classic Psychedelics: A Qualitative Study of Help-Seeking Experiences

**DOI:** 10.64898/2026.05.23.26353427

**Authors:** Lisa Maria Jöbstl, Bente Lubahn, Ebru Kaya, Georg Leistenschneider, Marija Franka Žuljević, Thomas G. Riemer, Dario Jalilzadeh Masah, Derin Marbin, Barbara Stöckigt, Tomislav Majić

**Affiliations:** Department of Psychiatry and Neurosciences, Charité – Universitätsmedizin Berlin, Corporate member of Freie Universität Berlin and Humboldt-Universität zu Berlin, Berlin, Germany; Department of Medical Humanities, University of Split School of Medicine, Split, Croatia; Institute of Clinical Pharmacology and Toxicology, Charité – Universitätsmedizin Berlin, Corporate member of Freie Universität Berlin and Humboldt-Universität zu Berlin, and Berlin Institute of Health, Berlin, Germany; Institute of Social Medicine, Epidemiology and Health Economics, Charité – Universitätsmedizin Berlin, Corporate member of Freie Universität Berlin and Humboldt-Universität zu Berlin, Berlin, Germany; Institute for General Practice and Interprofessional Care, University Hospital Tübingen, Tübingen, Germany; Robert Bosch Center for Integrative Medicine and Health, Bosch Health Campus, Stuttgart, Germany

**Author notes:** **Corresponding Author:** Dr. med. Tomislav Majić, Psychiatrische Universitätsklinik der Charité im St Hedwig Krankenhaus Große Hamburger Straße 5-11 10115 Berlin, Germany. contributed equally (Barbara Stöckigt, Tomislav Majić).

**Keywords:** psychedelic, HPPD, depersonalization, derealization, harm reduction

## Abstract

**Background:** While growing enthusiasm for the therapeutic potential of classic psychedelics has led to a rise in non-clinical use, attention to persisting adverse effects has emerged with delay. A subset of individuals reports persisting complications such as hallucinogen persisting perception disorder (HPPD), depersonalization/derealization disorder (DDD), anxiety and depression. Yet few medical services are equipped to address these complications.

**Aims:** This qualitative study examines how societal, medical, and media discourses shape the experiences of individuals with persisting psychedelic-related complications, focusing on help-seeking trajectories.

**Methods:** Thirteen semi-structured interviews with adults experiencing persisting psychedelic-related psychological symptoms (four women, nine men, age 19–49 years; HPPD (n = 10), DDD (n = 6), depression (n = 1), and anxiety (n = 1)) were conducted within a larger study on these complications. Data were analysed using reflexive thematic analysis. Reporting followed the COREQ guidelines.

**Results:** Three interrelated themes emerged: (1) The dissonance between expectation and harm — idealised media and scientific portrayals of psychedelics shaped initial use and complicated recognition of adverse outcomes; (2) Stigma, silence, and self-blame — prohibitionist discourse and internalised shame significantly inhibited help-seeking; and (3) Between systemic absence and self-organised support — participants encountered clinical unpreparedness and epistemic dismissal, which often led them to rely on online peer communities and self-management strategies. Positive clinical encounters, characterised by professional expertise and nonjudgmental engagement, were experienced as helpful.

**Conclusions:** Adequate clinical and conceptual frameworks for persisting psychedelic-related complications are lacking. An interdisciplinary, experience-informed approach integrating realistic risk communication, clinician training, and destigmatisation is required to support affected individuals.

## Introduction

The history of human research in classic psychedelics has been characterized by waves of polarized debates, ranging from demonization to over-enthusiastic claims regarding their healing properties. While the ritual use of substances like psilocybin and mescaline in indigenous traditions spans millennia, Western scientific engagement with these compounds only emerged in the early twentieth century (Beringer, 1927). With the discovery of the psychoactive properties of lysergic acid diethylamide (LSD) in 1942 (Hofmann, 2010), LSD and related psychedelics became subjects of clinical investigation, with early studies exploring their potential in psychotherapy and addiction treatment (Belouin & Henningfield, 2018; Johnson et al. 2014). However, as LSD use spread beyond laboratories into the counterculture of the 1960s, the social and political climate shifted dramatically (Dyck, 2009). Psychedelics became entangled with moral panic, politicization, and state control (Dyck, 2005, 2023) — culminating in their criminalization under the U.S. *Controlled Substances Act* of 1971, subsequently resulting in a standstill of research for decades.

Since the late 2000s, scientific interest in the therapeutic potential of psychedelics has resumed (Grob et al., 2010; Moreno et al., 2006). Clinical trials have delivered promising results in the treatment of several psychiatric disorders, including depression and anxiety in life-threatening illness (Griffiths et al., 2016), major depression (Goodwin et al., 2022), and substance-use disorders (Bogenschutz et al., 2015, 2022; Klaus et al., 2025). Alongside this re-emerging therapeutic optimism, public discourse has shifted again towards idealizing psychedelics as near-miraculous panacea for any indication, agents of healing and transformation (Evans et al., 2023; Meling et al., 2024). This enthusiasm reflects both early countercultural narratives (Metzner et al., 2008) and more recent waves of scientific popularization, often associated with what has been termed the “Pollan effect” following the wide reception of Michael Pollan’s book *How to Change Your Mind* (Pollan, 2018). While such narratives have contributed to destigmatization and renewed research interest (Carhart-Harris & Goodwin, 2017; Nutt et al., 2013), they may also risk narrowing public and clinical attention toward predominantly positive outcome trajectories (Schlag et al., 2022), obscuring the small but significant subset of individuals who experience persisting psychedelic-related complications (Evans et al., 2023).

In parallel with expanding clinical trials and public interest, the use of psychedelics has increased outside regulated therapeutic settings, including recreational, self-exploratory, and informal therapeutic contexts (Baxter et al., 2024). Recent reports from emergency and acute care settings suggest a corresponding rise in presentations related to adverse psychological reactions following psychedelic use, particularly among younger populations and individuals with pre-existing vulnerabilities (Bremler et al., 2023; European Union Drugs Agency, 2025; Evans et al., 2023; Kopra et al., 2022; Myran et al., 2024; Palamar & Le, 2023).

In this respect, the contemporary discourse surrounding psychedelics echoes a broader historical pattern in the medical use of psychoactive substances, where therapeutic optimism has frequently outpaced the systematic evaluation of potential harms (Healy, 2002; Rasmussen, 2008). Similar trajectories can be observed in the medical adoption of other substances, such as cocaine in late nineteenth-century psychiatry (Freud, 1884), the widespread promotion of opioid-containing cough remedies in the early twentieth century (Courtwright, 2001; Musto, 1999), the extensive and largely undifferentiated prescription of benzodiazepines between the 1960s and 1980s (Healy, 2004; Lader, 2011), and, more recently, the clinical and public health consequences associated with long-term opioid analgesic use (Healy, 2002), as well as the overly optimistic claims about the therapeutic potential of cannabinoids (Englund et al., 2017; Hall et al., 2019). Against this backdrop, emerging calls for more differentiated monitoring of risks, adverse outcomes, and vulnerable populations in the context of psychedelic use can be understood as part of a recurring pattern in the integration of psychoactive substances into medical and public discourse (Aday et al., 2020; Schlag et al., 2022). This underscores the need for systematic, empirically grounded approaches to identifying, classifying, and addressing both intended therapeutic effects and unintended psychological complications (Bremler et al., 2023; Majić et al., 2026).

Long-term psychedelic-related complications include psychedelic-related psychosis (Bröcker et al., 2026) persistent anxiety (Viljoen & Betzler, 2024), derealization and depersonalization disorder (DDD) (Jalilzadeh-Masah et al., 2025; Michal, 2025), and Hallucinogen Persisting Perception Disorder (HPPD) (Martinotti et al. 2018; Žuljević & Majić, 2025). HPPD is recognized in the *DSM-5* (code 292.89) (5th ed.; American Psychiatric Association [APA], 2022) as a disorder characterized by enduring visual disturbances reminiscent of those experienced during acute intoxication (Hermle et al., 2012). Symptoms can persist for weeks, months, or even years and may severely impair daily functioning (Halpern et al., 2016; Hermle et al., 2008, 2012; Leistenschneider et al., 2024; Žuljević & Majić, 2025). The prevalence of persisting psychedelic-related complications remains insufficiently characterized, as systematic epidemiological data are scarce (Simonsson et al., 2023) and underreporting is likely due to stigma, retrospective recall bias, and diagnostic uncertainty (Argyri et al., 2025; Carbonaro et al., 2016). Recent qualitative research underscores the complexity of these phenomena, linking adverse outcomes to an interplay of pharmacological, psychological, and contextual factors — summarized in the “set, setting, and substance” framework (Bremler et al., 2023; Metzner et al., 2008; Zinberg, 1984).

Individuals with persisting psychological psychedelic-related complications remain largely underrepresented in clinical and public discourse, leaving limited conceptual space within which their experiences can be described or understood. This epistemic imbalance represents not only a research gap but also a social and ethical blind spot. While clinical and neurobiological research on psychedelics flourishes, few studies address how individuals experiencing related complications navigate social and medical systems, and studies offer limited insight into how affected individuals interpret their experiences or encounter (mis)recognition within therapeutic and societal contexts. This study seeks to address this gap through an interpretive, qualitative lens. Specifically, it explores how individuals who experience long-term psychedelic-related complications describe their encounters with medical and social support systems, and how broader cultural narratives about psychedelics shape these experiences. By doing this, the study aims to (1) reconstruct how societal and medical narratives shape how individuals with psychedelic-related complications understand their conditions, (2) examine how these dynamics influence their help-seeking trajectories, and (3) identify implications for clinical and public discourse. This question acknowledges that experiences of illness are not merely biological events but are embedded within social narratives, expectations, and structures of meaning. By foregrounding *assumptions* (*Vorannahmen*), the study draws on hermeneutic and social-constructionist traditions (Gadamer, 1967; Berger & Luckmann, 1967), emphasizing how preconceptions — shaped by culture, media, and medicine — mediate both the interpretation of distress and access to care.

## Methods

### Study Design and Context

This study forms part of a broader research project investigating persisting psychedelic-related complications (*PPaPS study*) at Charité University Medicine in Berlin through both quantitative and qualitative methods. The present qualitative sub-study employed a reflexive thematic analysis (Braun & Clarke, 2006; Braun & Clarke, 2024) to explore subjective experiences of individuals with diagnosed persisting psychedelic-related complications, focusing on how social and medical assumptions shaped their help-seeking trajectories. This qualitative study was informed by established qualitative reporting frameworks to enhance methodological transparency and coherence, including the *Consolidated Criteria for Reporting Qualitative Research* (COREQ) (Tong et al., 2007) and recent values-based reporting guidelines for qualitative inquiry (Braun & Clarke, 2024), which advocate for methodologically congruent and reflexively transparent reporting practices in thematic research.

### Participants

Participants were recruited from the database of the larger, abovementioned study, which includes adults reporting persisting psychological symptoms temporally related to the use of psychedelics. Eligible participants had to be 18 years or older, fluent in German, and were diagnosed with psychological symptoms (e.g., anxiety, derealization, depersonalization, or perceptual disturbances) persisting for at least 72 hours beyond the acute effects of a psychedelic experience (“index experience”), which they retrospectively identified as the onset of their current difficulties.

### Sample Size Rationale

Thirteen of 25 eligible individuals agreed to participate (response rate: 52%). Reasons for non-participation were not assessed, as no feedback was received from non-responders. The final sample size was guided by the principle of thematic sufficiency as proposed by Braun and Clarke (2019), referring to the point at which the dataset is sufficiently rich to support a detailed and nuanced analysis of the research question. During data collection and analysis, recruitment was discontinued once additional interviews did not yield substantially new themes but rather deepened and refined existing patterns, indicating that sufficient conceptual depth had been achieved.

In the present study, interviews ranged from 20 to 68 minutes and generated substantive, detailed accounts across all four thematic domains of the interview guide. Across the thirteen interviews, the analytic team identified consistent patterns of meaning while also attending to variation and divergence between accounts. No new themes of conceptual significance emerged in the final three interviews, and the dataset was judged analytically sufficient to support the three themes reported. This assessment is consistent with published guidance suggesting that six to twelve interviews typically yield sufficient depth for a focused qualitative study (Braun and Clarke, 2019; Malterud et al., 2015), though the appropriate number remains context dependent. The relatively focused research question — concerning help-seeking experiences within a specific, diagnostically defined population — further supports the adequacy of this sample size.

### Data Collection

Interviews were conducted via encrypted video calls (ZOOM®) between February and August 2025 by the same researcher (L.M.J.) to ensure consistency in data collection. Each interview lasted 20 to 68 minutes. The semi-structured format allowed for open narratives while ensuring coverage of key themes.

The interview guide comprised four thematic blocks:

1. Personal attitudes and experiences with psychedelics — including context, motivation, and the index experience, i. e., the substance experience that preceded symptom onset.
2. Medical help-seeking experiences — focusing on interactions with healthcare professionals and perceptions of medical understanding or dismissal.
3. Social environment and narratives — exploring societal narratives, family reactions, and peer attitudes toward psychedelics.
4. Reflections on discourse and self-perception — examining how public portrayals of psychedelics influenced participants’ interpretations of their experiences and help-seeking.

Interviews encouraged narrative flow, with minimal interruption. Participants were invited at the end to add reflections or information they felt had not been addressed. Audio was transcribed verbatim using Whisper.ai and manually verified for accuracy. Identifying details were pseudonymized, and all recordings were deleted after transcription in compliance with ethical guidelines explained below.

### Data Analysis

Data were analysed following Braun and Clarke’s (2006) six-phase framework for reflexive thematic analysis: (1) Familiarization - transcripts were read repeatedly to identify recurrent meanings and patterns; (2) Initial coding - data segments relevant to the research question were coded inductively, capturing both semantic and latent meanings; (3) Searching for themes — codes were clustered across interviews into potential themes; (4) Reviewing themes — themes were refined to ensure internal coherence and distinctiveness; (5) Defining and naming themes — thematic labels were developed to capture the essence of each pattern; and (6) Producing the report — themes were contextualized within theoretical and empirical literature to interpret how cultural narratives intersected with personal experiences.

The coding process was conducted with the coding software f4analyse. Coding and thematic development were discussed iteratively within the PPaPS research team (including L.M.J. and senior researchers involved in the study) to enhance reflexivity, intersubjectivity, and interpretive rigor. Regular team discussions were used to review coding decisions and emerging themes. Discrepancies in interpretation were addressed through reflexive dialogue and resolved by consensus, ensuring that divergent perspectives were critically examined and integrated where appropriate. The analysis prioritized conceptual depth and experiential coherence rather than frequency counts, consistent with the interpretive nature of the study.

### Ethical Considerations

The study adhered to the ethical standards of the German Society for Psychology (Deutsche Gesellschaft für Psychologie [DGPs], 2022) and the Declaration of Helsinki (World Medical Association [WMA], 2013). All participants provided written informed consent as part of the PPaPS parent study and reconfirmed verbal consent for interview recording and analysis. Participation was voluntary and could be withdrawn at any time without consequence. Pseudonymization ensured confidentiality, and participants were informed of available support resources in case of emotional distress.

### Reflexivity and Positionality

Reflexive thematic analysis, as conceptualised by Braun and Clarke (2006, 2024), positions the researcher not as a neutral conduit of data but as an active participant in the production of meaning. In accordance with this epistemological stance, we acknowledge the following positionalities as relevant to the design, conduct, and interpretation of the present study. The institutional and professional context of Charité University Medicine Berlin carries implicit orientations: familiarity with psychiatric nosology may have foregrounded diagnostic framings of participants’ experiences, while the team’s broader investment in the systematic study of psychedelic-related complications (via the PPaPS project) reflects a commitment to legitimising this population as a clinical concern. We were alert to the possibility that this framing could inadvertently pathologise experiences that some participants themselves did not narrate in clinical terms. Some members of the research team have prior personal or professional engagement with psychedelic substances or psychedelic-assisted therapy research. This experience was understood as both a potential sensitising resource — enabling recognition of culturally specific narratives — and a source of interpretive bias requiring ongoing scrutiny. The primary researcher (L.M.J.) has formal training in clinical psychology with a strong focus on research methodology, including qualitative approaches. The dominant discourse within current psychedelic research is strongly optimistic. We were conscious that this context could create subtle pressure to frame adverse experiences as exceptional or self-limiting. Throughout analysis, we actively worked against this tendency by attending to experiences that resisted integration or recovery narratives.

Reflexivity was practised at multiple levels. Coding and thematic development were conducted iteratively and discussed within the research team to surface divergent interpretations and challenge premature closure. Assumptions were documented in reflective memos throughout the analytic process. Participants were not involved in member-checking of the final themes; however, the interview guide invited participants to reflect on and add to their accounts at the close of each session, providing a degree of respondent input into data completeness. We do not claim that these practices eliminate interpretive influence, but rather that they render it transparent and subject to critical scrutiny — consistent with the values-based framework for qualitative reporting advocated by Braun and Clarke (2024).

## Results

From 25 eligible individuals, 13 agreed to participate in semi-structured interviews (four women and nine men; all cisgender; age range: 19–49 years). Participants represented diverse educational and occupational backgrounds, reflecting a cross-section of those affected by long-term adverse psychedelic experiences. Some had prior psychiatric histories (e.g. depression, anxiety, trauma), and several explicitly linked their psychedelic experiences to self-experimentation motivated by therapeutic or spiritual goals (see *Table 1*). Pseudonyms are used for all participants.

**Table 1.** Sociodemographic characteristics, interview details, and psychedelic-related clinical profiles of participants (N = 13), listed in chronological order of interviews. The index experience refers to the psychedelic exposure retrospectively identified by each participant as temporally and subjectively linked to the onset of persisting psychological complications. Diagnoses were established as part of the PPaPS parent study at Charité University Medicine Berlin.

| Pseudonym | Gender | Age Range (at Interview) | Education | Duration of Interview (min) | Index Experience (IE)* |  |  | Psychedelic-related diagnoses |
| --- | --- | --- | --- | --- | --- | --- | --- | --- |
|  |  |  |  |  | Time since IE | Setting of IE | Substance(s) used |  |
| Paul | m. | 19-29 | High school | 26 | 2 years | With friends at home | LSD | HPPD, DDD |
| Max | m. | 19-29 | A-Levels | 28 | A few months | With friends at home | LSD | HPPD, DDD |
| Andrew | m. | 19-29 | A-Levels | 20 | 6 years | With a friend at home | Psilocybin | HPPD, Depression |
| Roman | m. | 30-39 | Masters | 25 | 4 years | Alone at home | LSD, Psilocybin (Microdosing) | DDD |
| Marie | f. | 19-29 | A-Levels | 68 | A few months | With friends outside in the forest and city | Psilocybin | Anxiety Disorder, DDD |
| Chris | m. | 40-49 | High school | 26 | 2 years | Alone at home | LSD | HPPD |
| Manuel | m. | 19-29 | High school | 27 | 4 years | With friends at home | Psilocybin | HPPD |
| Maik | m. | 40-49 | Masters | 31 | 25 years | Party/festival context | LSD | HPPD |
| Julia | f. | 30-39 | A-Levels | 65 | 2 years | Alone at home | LSD | HPPD |
| Tobias | m. | 19-29 | A-Levels | 21 | 2 years | Party/festival context | LSD + Cannabis | DDD |
| Lea | f. | 40-49 | A-Levels | 36 | 30 years | With friends at home | LSD | HPPD |
| Selina | f. | 30-39 | Masters | 28 | 8 years | Party/festival context | Psilocybin | HPPD, DDD |
| Luca | m. | 30-39 | Masters | 38 | 10 years and 2.5 years | Party/festival context | LSD | HPPD |

\***Index experience** = The psychedelic exposure retrospectively identified by participants as temporally and subjectively linked to the emergence of persisting psychological complications.
\*\***Diagnoses:** HPPD = Hallucinogen Persisting Perception Disorder, DDD = Depersonalization-Derealization Disorder

This study examined how societal and medical assumptions about psychedelics shape the lived experiences and help-seeking behaviour of individuals with persisting psychological symptoms associated with psychedelic use. Three interrelated processes emerged. Together, these processes delineate an interpretive trajectory from attraction to alienation:

1. The dissonance between expectation and harm — the influence of cultural, therapeutic, and media-based narratives on initial engagement with psychedelics and subsequent interpretations of adverse outcomes.
2. Stigma, silence, and self-blame — the entanglement of moral judgment and illegality in shaping help-seeking trajectories.
3. Between systemic absence and self-organized support — the negotiation of care and meaning within an epistemically underdefined condition.

Following sections elaborate the themes in detail whereas the third theme includes three analytic subthemes (see chapter 3.3.). The analysis draws on exemplary participant quotations, which were translated from German. The interview excerpts have been selectively abridged for readability while preserving their analytic integrity. Quotations are referenced by pseudonym and line number (l.) from the verbatim transcript. A comprehensive thematic overview including original German excerpts for all illustrative quotations is provided in *Supplementary Table S1*, supporting methodological transparency and translation verification. In order to maintain analytic clarity and narrative flow, the main text focuses on thematic synthesis, while additional verbatim excerpts are presented in the table to ensure transparency and traceability of interpretations.

### 1. The Dissonance Between Expectation and Harm

The analysis indicates that participants’ initial motivations for psychedelic use were closely shaped by prior expectations derived from dominant cultural and therapeutic narratives. Psychedelics were frequently framed as “tools of healing”, “gateways to self-knowledge”, or natural alternatives to conventional psychiatry. More than half of the participants explicitly referred to podcasts, documentaries, and online articles portraying substances such as psilocybin or LSD as promising treatments for depression, trauma, or anxiety. These media representations, often perceived as conveying scientific legitimacy, informed participants’ decisions to engage with psychedelics — typically framed as therapeutic or spiritual practices rather than recreational drug use. Importantly, these expectations continued to shape how participants interpreted emerging symptoms and influenced their subsequent help-seeking trajectories.

Roman described how media portrayals of psychedelic-assisted therapy fuelled hope that psychedelics might alleviate suffering: “*I came across it through some documentary — about all the things it can help with. But it has its risks too. That’s something I unfortunately found out the hard way.*” (Roman, l. 4–9). His account illustrates a widespread ambivalence: psychedelics were perceived as transformative and healing, yet their risks were less visible or understood until encountered personally. Similarly, Julia reported fascination with idealized and mystical representations circulating in popular culture: “*Psychonauts, seeing machine elves− I wanted to try that for myself. But then, of course, it was the complete opposite [and I experienced negative effects].*” (Julia, l. 12–24). Here, media-driven imagery contributed to a narrative of adventure and self-discovery, which strongly shaped expectations — making the negative turn of events appear particularly surprising and challenging to integrate. The findings indicate that promising media narratives and popular therapeutic claims influenced not only many participants’ initial motivation to use psychedelics but also shaped their subsequent interpretation of emerging difficulties.

Chris further emphasized how shifting public discourse reframed psychedelics as a legitimate tool for growth and well-being: “*In the media it’s being spread everywhere right now − great remedy, completely harmless, it can’t be that dangerous, maybe a new impulse for my personal development.*” (Chris, l. 34–43). This reflects a cultural normalization and risk-minimization that lowers thresholds for use and embeds psychedelic experiences into broader narratives of self-optimization — illustrating a pattern shared by several participants. For Marie, such cultural ideals were incorporated into identity and future aspirations: “*Since I was 13, I already knew that I wanted to try LSD or mushrooms at some point. The last [psychedelic experience] was with mushrooms. And it went very, very badly.*” (Marie, l. 8–15). Her account demonstrates how psychedelic use may become biographically anticipated and socially legitimized, even outside traditional subcultures.

Across more than half of the interviews, participants described how exposure to media representations, popular science accounts, and therapeutic framing — embedded within the broader discourse of the “psychedelic renaissance” — shaped their expectations and acted as a key influence on their decisions to initiate psychedelic use, while also informing how subsequent adverse experiences were interpreted. In contrast, approximately one third of participants reported primarily recreational motivations. Their psychedelic use was situated within party, festival, or peer-group contexts and framed as exploratory, social, or pleasure- oriented rather than explicitly therapeutic. For this group, expectations were less shaped by clinical or self-optimization narratives and more by curiosity and the pursuit of altered states within leisure settings.

### 2. Stigma, Silence, and Self-Blame

After the onset of persisting psychedelic-related psychological complications, societal and internalized stigma surrounding psychedelic use significantly inhibited help-seeking among the great majority of the interviewed individuals. The legacy of prohibitionist and moralistic drug discourse persisted in subtle and overt forms, shaping not only social reactions but mostly participants’ self-perceptions and interactions with healthcare systems. Psychedelics were widely associated with illegality, irresponsibility, and irrationality, informing expectations of social disapproval. Julia similarly linked her difficulty seeking help to early learned moral narratives: “*Drugs are bad. Everything that is illegal is definitely bad.*” (Julia, l. 87–90).

Participants very often anticipated ridicule and moral judgment from others, which led many to conceal their experiences. Roman described a cultural discourse arising for him after the onset of his psychedelic-associated symptoms that oscillates between danger and absurdity: *“[The people around me think:] That stuff gives you psychosis and makes you think you’re a banana — just some hippie, completely embarrassing.*” (Roman, l. 108–123). Such cultural framings were not merely external but deeply internalized, producing shame and self-blame. Lea, consistent with most accounts across the sample, articulated this dynamic clearly: “I was absolutely ashamed of myself. Completely reckless, crazy, irresponsible — being branded as such.” (Lea, l. 85–104). As a result, many participants struggled to disclose their symptoms even to close contacts. Paul emphasized his caution: “*You have to be very careful about who you tell something like that. I also thought that people would have talked about it behind my back somehow. And that’s what I wanted to avoid.*” (Paul, l. 62–75). Self-stigma also emerged as a barrier to recognizing one’s own legitimacy to seek care. Participants frequently described their suffering as self-inflicted, making them feel undeserving of support: “*I think I was ashamed that I had caused it myself.*” (Luca, l. 256–257). Some participants also pointed to the broader stigma surrounding mental health as a compounding factor.

### 3. Between Systemic Absence and Self-Organized Support

Participants’ accounts reveal a field of tension between systemic unpreparedness and the possibility of meaningful care. While the majority encountered clinical structures that offered few interpretive frameworks or therapeutic pathways for their experiences — frequently resulting in feelings of not being understood and withdrawal from formal support — a notable portion of participants also described episodes of helpful, informed clinical engagement that proved pivotal in their recovery trajectories. Alongside these institutional encounters, individuals actively negotiated this void by building informal networks, seeking information independently, and developing personal coping strategies.

Three interconnected subthemes emerged:

1. The absence of professional understanding and clinical recognition
2. The emergence of self-organized support and alternative coping practices
3. Helpful clinical support as a restorative resource

Taken together, the following theme highlights a complex field of tension between systemic barriers, subjective meaning-making, and moments of genuine understanding within care contexts.

#### 3.1 The absence of professional understanding and clinical recognition

> *“No one can really help me.”* (Lea, l. 83–95)

This subtheme captures participants’ reports of clinical unpreparedness and dismissal, which frequently led to feelings of not being taken seriously and lacking a legitimate place for their experiences within the healthcare system. Selina’s account illustrates this foundational issue on a conversation with her psychiatrist: “*But then he said, okay, I’ll have to bring this up at the next conference. So he simply had no knowledge of it at all.*” (Selina, l. 49–51). Participants consistently encountered a lack of knowledge about psychedelic-associated conditions, which limited diagnostic guidance and undermined trust. Marie’s experience exemplifies how moralizing responses replaced clinical engagement: “*He [the doctor] had rather little time and didn’t really know much about it. [He said:] ‘One shouldn’t take things like that… Magic mushrooms — what even is that?’*” (Marie, l. 256–293). A perceived absence of expertise often led to premature diagnostic assumptions and foreclosed meaningful dialogue. Manuel explained how this expectation undermined hope and shaped self-censorship: “*None of them are really specialised in that sort of thing [psychedelics] either, so I didn’t have much hope there.*” (Manuel, l. 65–74). Roman’s account demonstrates representative for most participants how the refusal to recognize psychedelic involvement as clinically relevant intensified suffering and induced profound insecurity: “*I had no idea what was happening [to me]. There was a lot of [medication] experimentation. I didn’t know that people could suffer so much.*” (Roman, l. 133–173). Here, epistemic injustice became emotionally destabilizing: his knowledge was discounted while standard psychiatric interventions worsened symptoms. Julia’s statement reflects the invalidating impact of being misdiagnosed in contradiction to one’s own cognitive clarity, escalating into feelings of loss of agency: “*Nobody is listening to me. Help, help.*” (Julia, l. 486–487).

Among others, Tobias highlighted barriers both structural and stigmatizing - long waiting lists, excessive administrative burden, and fear of judgment regarding drug use:

> *You spend hours on the phone, you write emails — and all of that while you’re in a state where you feel absolutely awful, and every social interaction drains you completely. And on top of that, with the thought in the back of your mind: how openly can I even talk about drug use here at all. All of that took an enormous amount of energy.* (Tobias, l. 215–225)

Finally, participants expressed urgent calls for improved provider training and system-wide awareness: “*And I think it would be really great if there were a bit more knowledge out there about how to actually treat this.*” (Marie, l. 490-492).

#### 3.2 The emergence of self-organized support and alternative coping practices

Repeated encounters with disbelief or dismissal in clinical settings prompted many participants to shift their search for support away from institutional care and toward self-organized information-seeking and coping strategies. Online communities — particularly platforms such as Reddit — often emerged as central sources of orientation when formal services failed to provide explanations or actionable support. For some participants, engagement with online communities represented the first encounter with clinical framings such as HPPD. As Max noted: “*The only real resource I could find anything out through was Reddit. A very large community that’s quite active and posts about it — to try to understand it a little.*” (Max, l. 53–55). Some participants emphasized that shared experiences in digital spaces offered both practical knowledge and emotional validation. For some, this shift involved accessing unfamiliar platforms or peer groups in moments of acute distress. Marie recounted reading narratives from an English-speaking Facebook community: “*And reading what was posted there — I have to say, my case is probably not that bad.*” (Marie, l. 347–358). Here, comparative peer narratives helped recontextualize her symptoms and reduce fear, demonstrating the normalizing and stabilizing effects of online support networks.

Several participants also described active self-management strategies, ranging from dietary supplements to exercise and mindfulness practices. Paul reported experimenting with anxiolytic options he identified through online reading: “*And then I read about what you can do for anxiety. You can take valerian, then GABA — and that helped even more.*” (Paul, l. 263–270). Lifestyle changes were similarly described by Max - “*started exercising and eating more healthily*” (l. 74–75) — and by Luca — “*did a lot of yoga*” (l. 143–144). These narratives highlight how individuals turned toward proactive health behaviours to regain stability in the absence of sufficient medical guidance.

Such self-directed engagement also enabled the development of personal interpretive and integration frameworks for some participants, as illustrated in Manuel’s reflective sense-making: “*It’s quite possible that the LSD made me enormously susceptible to those relationship dynamics [with my ex-partner]. That would be my theory about it.*” (Manuel, l. 221–224). His account demonstrates how participants sought coherence and control by constructing their own explanatory models, integrating psychedelic experiences into broader biographical and relational contexts. Overall, more than half of the participants described turning to alternative avenues for information, reassurance, and healing — pathways that provided room for individualized recovery processes beyond traditional clinical structures.

#### 3.3 Helpful clinical support as a restorative resource

For many participants, access to helpful care depended on their own efforts to locate knowledgeable professionals or specialized clinics, underscoring systemic gaps in routine mental-health provision. When such support was found, participants emphasized two core qualities: (1) professional expertise in psychedelic-related phenomena, and (2) a non-judgmental therapeutic stance that acknowledged their subjective experiences as real and meaningful.

Manuel described, consistent with accounts across the sample, how professional reassurance reduced fear and reframed his symptoms: “*And they also reassured me that I had not sustained any permanent damage because of it. And I hadn’t broken anything with the LSD. I didn’t go crazy because of it or anything like that.*” (Manuel, l. 68–73). Reflecting on his psychiatric encounter, he emphasized the importance of being both understood and taken seriously: “*Yes, [the experience with my psychiatrist was] definitely positive, because he really knows his stuff. And then you somehow felt seen and understood.*” (Manuel, l. 107–114). Similarly, Julia valued a therapeutic setting in which even unusual perceptual experiences could be shared without immediate dismissal: *“[I told the psychiatrist] I took the LSD. My voice sounds strange to me. I can see colours on the wall. And it was not blocked, but… everything was taken in.*” (Julia, l. 559–563). Positive encounters were predominantly described as turning points in the help-seeking process. Roman explained how clear diagnostic feedback reduced existential anxiety and enabled symptom interpretation:

> *But at some point, I simply couldn’t go on anymore. Then I was admitted to the clinic. And then I said to myself, okay, come on, give it one more chance and go to this [psychiatrist]. And it really only took four, five hours with her before she immediately recognised it — ah, okay, that’s what’s going on with this young man. And then I realised it myself too: okay, this is just derealisation, and I’m not dying, I’m not getting a psychosis, or anything like that.* (Roman, l. 36–41)

Likewise, Tobias emphasized that a judgment-free clinical space enabled disclosure of psychedelic use: “*Yes, okay, then I’ll be able to talk freely about having used [substances]*.” (Tobias, l. 205–206). Across accounts, recognition and validation were experienced as therapeutically powerful in themselves. Helpful clinical interactions enabled participants to stabilize overwhelming experiences, reinterpret symptoms with reduced fear, and regain a sense of psychological coherence. Importantly, these encounters counteracted prior experiences of disbelief and helped restore trust in professional care.

## Discussion

In our study investigating help-seeking experiences in individuals suffering from psychedelic-related complications, three thematic clusters were identified that can be understood as reflecting a broadly sequential pattern in participants’ retrospective accounts of their experiences: (1) The dissonance between expectation and harm (first phase): A positive interpretative framework was established that encouraged use and shaped expectations of positive psychedelic experiences associated with the idea of healing. This framework was destabilized when participants reported persisting psychological complications that stood in contrast to their initial expectations of benefit. (2) Stigma, silence and self-blame (second phase): Optimism was replaced with alienation, as individuals encountered internalized and social judgment and medical invalidation, and (3) Between systemic absence and self-organized support (third phase): Participants developed pragmatic responses to institutional voids and regained a feeling of self-efficacy, control, and stability, alongside occasional experiences of helpful clinical engagement.

Across all themes runs a common thread: the absence of legitimized language. Participants oscillated between discourses of therapy, pathology, and spirituality without finding a stable identity category that encompassed their reality. Their experiences expose a gap between the promises of the psychedelic renaissance (Meling et al., 2024; Schlag et al., 2022) and the lived consequences of its exclusions. The sequential trajectory identified in this analysis aligns with emerging evidence that expectations and cultural framing play a constitutive role in shaping both beneficial and adverse psychedelic experiences (Hartogsohn, 2016). Research on placebo and nocebo mechanisms suggests that pre-existing beliefs can modulate subjective outcomes and symptom interpretation, particularly in interventions characterized by heightened suggestibility and emotional salience (Benedetti, 2014; Pronovost-Morgan et al., 2023). Within psychedelic contexts, set and setting have been shown to exert substantial influence over acute phenomenology and longer-term meaning-making processes (Hartogsohn, 2016, 2018). The present findings extend this literature by illustrating how these same mechanisms operate beyond the acute phase, shaping how persisting symptoms are interpreted, legitimized, or dismissed over time. Rather than constituting a purely individual coping trajectory, the three phases observed here appear embedded in broader socio-cultural narratives that structure both help-seeking behaviour and perceived access to care.

### The Ambivalence of the Psychedelic Renaissance

The results highlight a paradox at the heart of the contemporary discourse on psychedelics. While the so called “psychedelic renaissance” promotes connectedness with oneself and others (Schutt et al., 2024), compassion (Zeifman et al., 2025, *preprint*) and healing (Swoap, 2023), its public rhetoric often reproduces one-sided optimism (Meling et al., 2024). Participants’ accounts reveal how exposure to overly positive narratives — via media, online communities, or scientific popularization — created unrealistic expectations and hindered recognition of risk. This dynamic has been further amplified by popular science narratives — often referred to as the “Pollan Effect” (see introduction) that contributed to the mainstream normalization and idealization of psychedelic use (Meling et al., 2024, Schlag et al., 2022). Similar critiques have been raised by other authors (Argyri et al., 2025; Bremler et al., 2023; Evans et al., 2023) who caution against the uncritical celebration of psychedelics as panacea.

From a science communication perspective, this pattern reflects what has been described as a “hype cycle” in biomedical innovation, whereby early-stage findings are rapidly translated into public narratives of breakthrough and transformation, often outpacing clinical evidence and risk communication (Caulfield, 2018). Similar dynamics have been documented across the history of psychopharmacology: the overprescription of cocaine in late nineteenth-century medicine (Freud, 1884; Courtwright, 2001), the widespread and largely undifferentiated use of benzodiazepines from the 1960s onwards (Lader, 2011; Healy, 2004), the escalation of opioid analgesic prescribing from the 1990s (Musto, 1999; Courtwright, 2001), and, more recently, overly optimistic claims regarding the therapeutic potential of cannabinoids (Englund et al., 2017; Hall et al., 2019). In each case, initial therapeutic promise overshadowed the systematic investigation of adverse outcomes. In the case of psychedelics, it is important to consider that decades of prohibition not only restricted systematic research into both beneficial and adverse effects but also limited the development of balanced clinical and public discourse. As a result, the current psychedelic renaissance has emerged in a context where therapeutic promise has been more prominently foregrounded than potential risks, contributing to a tendency toward one-sided optimism. Participants’ accounts suggest that popular and semi-scientific representations do not merely inform public attitudes but actively structure experiential expectations, influencing how distress is recognized and whether it is interpreted as a medical issue, a personal shortcoming, or a temporary “integration challenge”. This finding resonates also with sociological models of medicalization, which emphasize that symptoms become clinically actionable only when they are culturally legible as legitimate forms of illness (Conrad, 2007).

### Stigma and Epistemic Injustice in Help-Seeking

Taken together, the idealization of psychedelics may inadvertently silence those whose experiences contradict the dominant success narrative. When individuals encountered persisting psychological complications, the stark contrast to their expectations often led to disappointment and confusion. The lack of available social scripts for adverse outcomes leaves affected individuals linguistically stranded — a condition Fricker (2007) defines as *Hermeneutical Injustice*. Participants’ suffering appeared to unfold within an interpretive vacuum: they possessed abundant narratives for healing but none for persistent dysfunction. When psychedelic-related complications emerged, such as anxiety, derealization, or visual disturbances, participants struggled to integrate these experiences into existing narrative frameworks. Without culturally recognized terms for their suffering, individuals struggle to narrate it even to themselves, reinforcing alienation.

Parallel to idealization runs persistent stigma rooted in the decades of prohibitionist discourse. Despite growing clinical acceptance, the association of psychedelics with illegality and deviance remains deeply ingrained (Nutt et al., 2013). This study shows that stigmatization operates on two levels: *socially* (fear of judgment, ridicule) and *institutionally* (dismissal by clinicians). The participants’ fear of disbelief mirrors findings in other marginalized illness groups, such as functional disorders or ME/CFS, where patients confront *Testimonial Injustice* — the systematic undervaluation of their credibility (Fricker, 2007). In the context of persisting psychedelic-related psychological complications, this injustice is intensified by cultural ambivalence: affected individuals are viewed simultaneously as reckless drug users and as anomalies within a narrative of healing.

The reluctance to disclose symptoms and the anticipation of disbelief observed among participants is consistent with broader research on help-seeking in stigmatized mental health conditions. Studies across psychiatric and substance-related contexts indicate that perceived moral judgment and diagnostic uncertainty are key predictors of delayed care and disengagement from services (Clement et al., 2014; Corrigan et al., 2014). In clinical encounters involving medically contested or poorly defined conditions, patients frequently report epistemic asymmetries in which experiential knowledge is subordinated to biomedical authority, contributing to frustration and withdrawal from formal care systems (Carel & Kidd, 2014; Werner & Malterud, 2003). The present findings suggest that psychedelic-related complications may occupy a similar “contested” clinical space, where uncertainty about aetiology and classification intersects with moralized views of drug use, thereby amplifying barriers to recognition and treatment. Addressing stigma thus requires not only destigmatizing drug use but legitimizing negative psychedelic outcomes as worthy of clinical attention.

Together, these accounts show that psychedelic-related mental health crises occupy a doubly stigmatized position — marked both by drug-related moral judgments and by persistent societal devaluation of psychological suffering. This dual stigma seems to foster secrecy, restrict communication, and frequently delay or prevent access to appropriate professional support. These dynamics may help explain why many participants remained silent or delayed professional help. Furthermore, it can be argued that, within a discourse that strongly associates psychedelics with healing and insight, distressing symptoms were often construed as personal failure rather than as reflections of systemic neglect or substance- and context-related risks, thereby contributing to self-blame and a privatization of suffering.

The reported institutional gaps can also be understood through the lens of translational lag between experimental psychopharmacology and routine clinical practice. While controlled trials of psychedelic-assisted therapies have expanded rapidly, the integration of associated risk profiles and adverse outcome management into psychiatric training and service provision has progressed more slowly (Johnson et al., 2008; Phelps, 2017). The present study contributes qualitative evidence that such structural limitations are not merely organizational but are experienced at the interpersonal level as misrecognition and therapeutic discontinuity.

### Structural Gaps and the Rise of Informal Care

Participants’ reliance on online communities and self-organized networks indicates a transformation of isolation into autonomy emerging out of a significant deficiency within formal mental-health care. Current psychiatric classifications recognize HPPD but neglect broader forms of persisting psychological symptoms, such as DDD or mood disturbance. The absence of diagnostic clarity translates into therapeutic uncertainty: patients are frequently told that their symptoms are “psychotic” or “imagined”, which may erode trust and perpetuate suffering (Majić et al., 2026; Bremler et al., 2023). This epistemic gap reflects a lag between rapid scientific innovation and clinical implementation. As psychedelic research re-enters mainstream psychiatry, it is imperative that adverse and persistent effects are not sidelined. Interdisciplinary training programs — linking psychopharmacology, psychotherapy, and sociology — could equip clinicians to respond to persisting psychedelic-related psychological complications with both scientific and empathic competence. Overall, this aspect underscores that self-help emerged as a necessary and adaptive response when formal support systems were unprepared or unresponsive which reflects a regained sense of control over symptoms among affected individuals and even fosters self-efficacy.

### Strengths and Limitations of the Study

This study has several strengths. First, it addresses a largely underexplored population: individuals reporting persisting psychedelic-related complications. While clinical and experimental research has predominantly focused on therapeutic efficacy and acute safety, the present study contributes empirical insight into post-acute and enduring adverse experiences that remain underrepresented in both research and public discourse. Second, the qualitative design allowed for an in-depth exploration of lived experience, meaning-making processes, and help-seeking trajectories that cannot be captured through standardized symptom measures alone. By applying reflexive thematic analysis, the study was able to identify broader interpretative patterns that illuminate how societal narratives, stigma, and institutional responses interact in shaping participants’ experiences. The inclusion of a detailed thematic table enhances analytic transparency and traceability. Third, the study contributes to harm-reduction–oriented discourse by differentiating between heterogeneous forms of persisting psychedelic-related complications rather than reducing them to a single diagnostic entity. This nuanced approach may inform future research, diagnostic refinement, and the development of tailored clinical support structures. These strengths must be considered alongside several limitations. The sample size was self-selecting, which limits generalizability. Additionally, the study relies on retrospective self-report, which may be influenced by recall bias and post hoc meaning-making processes.

A further limitation concerns the gender composition of the sample. All thirteen participants identified as cisgender, and the sample was predominantly male (nine of thirteen). The absence of transgender, non-binary, and gender-diverse participants limits the transferability of findings to these populations, who may experience distinct intersections of stigma — combining drug-related, mental health-related, and gender-minority stigma — that could substantially shape help-seeking trajectories in ways not captured here. It cannot be determined from the present data whether this absence reflects genuine underrepresentation of gender-diverse individuals among those experiencing persisting psychedelic-related complications, differential willingness to participate in clinical research, or structural barriers within the recruitment pathway. Future research should adopt targeted recruitment strategies to ensure the inclusion of gender-diverse voices in this field. Furthermore, the study focuses exclusively on patient perspectives and does not incorporate clinician viewpoints or independent diagnostic verification beyond reported diagnoses.

The gender distribution within the cisgender subsample also warrants consideration. With nine male and four female participants, women are underrepresented relative to their presence in the general population and, potentially, among those affected by persisting psychedelic-related complications. Whether this imbalance reflects a genuine sex- or gender-related difference in the incidence or phenomenology of such complications, differential patterns of help-seeking and clinical referral, or a recruitment artefact specific to this study population cannot be determined from the present data. Research on stigma and help-seeking in adjacent conditions — including substance-related disorders and medically contested illnesses — consistently documents that women and men differ in the pathways through which they access care and in the ways they narrate and legitimise distress (Corrigan et al., 2014; Werner & Malterud, 2003). It is therefore possible that the themes identified in this study capture primarily male-gendered modes of experience and help-seeking, and that women’s trajectories involve distinct barriers or protective factors not represented here. Future research should employ recruitment strategies designed to achieve greater gender balance and to examine whether the three themes identified generalize across gender groups.

Finally, as with all qualitative research, findings are interpretative and context bound. While analytic rigor was maintained through reflexive transparency and adherence to qualitative reporting standards, themes represent constructions emerging from researcher–participant interaction rather than fixed categories inherent in the data. Taken together, the study should be understood not as an epidemiological estimate of prevalence or causal inference, but as an exploratory contribution that highlights an emerging clinical and societal phenomenon requiring further systematic investigation.

### Outlook

Future research should examine how exposure to the psychedelic renaissance shapes mental health professionals’ openness or bias, how moral and legal framings of drug use affect empathy and decision-making, and how professionals navigate uncertainty when faced with experiences beyond established biomedical paradigms. Further inquiries into the meaning of diagnosis, the conditions enabling empathic yet critical clinical engagement, and the motivations for entering this field are essential to advance both education and ethical practice in psychedelic psychiatry. Furthermore, future research should integrate phenomenological, clinical, and neurobiological perspectives to establish a multidimensional model of psychedelic-related complications. Quantitative surveys could assess prevalence and identify risk factors, while longitudinal designs may illuminate recovery trajectories. Importantly, participatory research involving individuals with lived experience could reduce epistemic hierarchies and foster more inclusive frameworks for understanding psychedelic harms.

To help address this gap, psychedelic research and policy could benefit from greater integration of biopsychosocial and hermeneutic perspectives that consider subjective accounts alongside neurobiological data. Clinical training may be strengthened by including education on psychedelic phenomenology, including potential adverse effects. Likewise, public discourse might profit from moving beyond binary framings of psychedelics as either “miracle medicines” or “dangerous drugs”, toward a more nuanced understanding of the diverse and context-dependent realities of psychedelic experiences.

### Conclusion

This study highlights that psychological difficulties following the use of classic psychedelics arise within a complex interaction of individual vulnerability, sociocultural expectations, and structural conditions of care. Participants’ accounts suggest that both contemporary enthusiasms surrounding psychedelic research and enduring moral stigma and prohibition shape how adverse or persistent experiences are understood, communicated, and addressed. Rather than acting in isolation, these forces jointly influence recognition of harm and access to support. The findings indicate that overly optimistic public narratives may contribute to unrealistic expectations, while stigma and limited clinical familiarity can impede timely help-seeking when experiences deviate from anticipated outcomes. In such contexts, individuals may struggle to find appropriate language or professional frameworks to articulate distress, particularly when symptoms do not align with established diagnostic categories. Importantly, these challenges coexist with well-documented therapeutic potentials of psychedelics under controlled conditions, underscoring the need for a balanced and differentiated perspective.

From a harm reduction perspective, the results point to the value of acknowledging the full spectrum of psychedelic experiences, including persisting adverse reactions, without framing them as either rare exceptions or inevitable outcomes. Clinical and educational approaches that emphasize informed consent, realistic risk communication, and openness toward subjective experience may help facilitate timely support and mitigate suffering. Increased professional awareness of psychedelic-related phenomenology, alongside a nonjudgmental stance toward substance use, could contribute to a more responsive and inclusive care. Overall, this study supports the integration of patient-centred perspectives into psychedelic research, clinical practice, and public discourse. A harm-reduction-oriented framework that recognizes both potential benefits and risks may be particularly well suited to navigate the evolving landscape of the use of psychedelics in therapy and beyond, while fostering ethical, compassionate, and evidence-informed responses to psychological distress.

## Supporting information

Supplemental: Coreq List

## Acknowledgements

The authors wish to thank all participants who generously shared their experiences for this study. Their openness and trust made this research possible.

## Statements and Declarations Ethical considerations

This study was approved by the Ethics Committee of *Charité – Universitätsmedizin Berlin*, approval number *EA2/233/23*. The study was conducted in accordance with the ethical standards of the German Society for Psychology (*Deutsche Gesellschaft für Psychologie*; DGPs, 2022) and the principles of the Declaration of Helsinki (World Medical Association, 2013). Ethics approval for the present qualitative sub-study was granted as part of the broader PPaPS study protocol.

## Consent to participate

All participants provided written informed consent as part of the PPaPS parent study prior to inclusion. Verbal consent for interview recording and analysis was reconfirmed at the commencement of each individual interview. Participation was entirely voluntary, and participants were explicitly informed of their right to withdraw at any time and without consequence.

## Consent for publication

Informed consent for the publication of anonymised interview material was obtained from all participants. All personally identifying details have been pseudonymised. Audio recordings were deleted following verbatim transcription in accordance with the ethical guidelines described above.

## Data availability

Given the sensitive and confidential nature of the qualitative data, raw interview transcripts are not publicly available. Anonymised excerpts are presented within the manuscript. De-identified data may be made available from the corresponding author upon reasonable request and subject to the conditions of the original ethics approval.

