## Supplemental Table 1 for "Persisting Psychological Complications Following the Use of Classic Psychedelics: A Qualitative Study of Help-Seeking Experiences"

### Supplementary Table S1

*Representative Participant Quotations by Theme, Subtheme and Subcategory*
*Original German excerpts with English translations and interpretive notes*

**Note.** This table provides the original German excerpts for all illustrative quotations used in the main text, together with additional exemplary quotes not included in the narrative. English translations appear in the main text; German originals are provided here to support translation verification and methodological transparency, in accordance with COREQ reporting standards. Excerpts have been selectively abridged for readability; abridgements are indicated by ellipses. Themes, subthemes and corresponding subcategories are marked with the same color to facilitate visual grouping within the table. The color coding has no analytical meaning. Participant names are pseudonyms. Z. = line number in transcript.

| Theme | Subcategories | German Original (participant, line ref.) | English Translation | Interpretive Note |
|---|---|---|---|---|
| **1. The Dissonance Between Expectation and Harm** | *Media-driven expectations* | *„Da gab es eine schöne Dokumentation, als wenn es halt quasi ein tolles Heilmittel wäre, völlig harmlos."* (Chris, Z. 34–43) | "There was a nice documentary, as if it were basically a great remedy, completely harmless." | Highlights how media narratives promoting psychedelics as benign and therapeutic shaped expectations prior to use. |
| | *Cultural fascination* | *„Psychonauten fand ich einen ganz tollen Begriff."* (Julia, Z. 12–24) | "I thought 'psychonauts' was a really great term." | Shows how cultural metaphors romanticise psychedelic use and construct an appealing identity framework prior to the index experience. |
| | *Lifelong normalisation* | *„Seit ich 13 Jahre alt bin, wusste ich schon, dass ich irgendwann mal LSD oder Pilze probieren möchte. Die letzte* [psychedelische *Erfahrung*] *war mit Pilzen. Und die ist sehr, sehr schlecht gegangen."* (Marie, Z. 8–15) | "Since I was 13 years old, I already knew that I wanted to try LSD or mushrooms at some point. The last [psychedelic experience] was with mushrooms. And it went very, very badly." | Demonstrates early incorporation of psychedelic narratives into identity and biographical life scripts. |
| | *Therapeutic hope* | *„Ich bin durch irgendeine Doku darauf aufmerksam geworden, gegen was das alles helfen kann. Aber hat halt auch seine Risiken. Das habe ich ja dann leider mitgekriegt."* (Roman, Z. 4–9) | "I came across it through some documentary — about all the things it can help with. But it has its risks too. That's something I unfortunately found out the hard way." | Illustrates the tension between therapeutic promise conveyed through popular science media and the unexpected adverse outcomes subsequently experienced. |
| | *Risk minimisation through public discourse* | *„In den Medien wird das gerade breitgetreten, tolles Heilmittel, völlig harmlos, so gefährlich kann es nicht sein, vielleicht einen neuen Impuls für meine persönliche Weiterentwicklung."* (Chris, Z. 34–43) | "In the media it's being spread everywhere right now — great remedy, completely harmless, it can't be that dangerous, maybe a new impulse for my personal development." | Reflects cultural normalisation and risk-minimisation that lowers the threshold for use and embeds psychedelic experiences within self-optimisation narratives. |
| | *Mystical / adventure framing* | *„Psychonauten, Maschinenelfen sehen, das probiere ich jetzt auch mal aus. Aber dann war es natürlich das komplette Gegenteil."* (Julia, Z. 12–24) | "Psychonauts, seeing machine elves — I wanted to try that for myself. But then it was of course the complete opposite (and I | Media-driven mystical imagery contributed to an adventure and self-discovery narrative that made the negative turn of events particularly |

| | | | | |
|---|---|---|---|---|
| | | | experienced negative effects)." | surprising and difficult to integrate. |
| **2. Stigma, Silence, and Self-Blame** | *Illegality and moral judgment* | *„Das ist ja verboten. Eine Droge zu nehmen, das ist einfach stigmatisiert.“* (Chris, Z. 109–110) | "It's forbidden after all. Taking a drug is simply stigmatised." | Reflects the moral-legal framing that shapes anticipated social judgement and deters disclosure of substance-related symptoms. |
| | *Early-learned moral narratives* | *„Drogen sind schlecht. Alles, was illegal ist, ist definitiv schlecht.“* (Julia, Z. 87–90) | "Drugs are bad. Everything that is illegal is definitely bad." | Links early socialisation into prohibitionist discourse with internalised barriers to help-seeking in adulthood. |
| | *Internalised shame* | *„Ich habe mich in Grund und Boden geschämt. Absolut leichtsinnig verrückt, unvernünftig — abgestempelt werden.“* (Lea, Z. 85–104) | "I was absolutely ashamed of myself. Completely reckless, crazy, irresponsible — being branded as such" | Shows deep internalisation of stigma — not merely anticipated from others but adopted as self-description — inhibiting both disclosure and help-seeking. |
| | *Fear of social consequences* | *„Man muss sehr doll aufpassen, wem man sowas erzählt. Ich denke auch, dass man hinter meinem Rücken dann darüber irgendwie geredet hätte. Und das habe ich vermeiden wollen.“* (Paul, Z. 62–75) | "You have to be very careful who you tell something like that to. I also think that people would have talked about it behind my back. And that is what I wanted to avoid." | Indicates concern about reputational harm and secondary gossip, leading to deliberate concealment of symptoms even from close contacts. |
| | *Self-blame* | *„Gefühl, da selber dran schuld zu sein.“* (Manuel, Z. 165–172) | "The feeling of being to blame for it myself." | Captures internalised responsibility narratives that diminish perceived legitimacy to seek professional care. |
| | *Self-blame — additional* | *„Ich glaube, ich habe mich geschämt, dass ich das selber verursacht habe.“* (Luca, Z. 256–257) | "I think I was ashamed that I had caused it myself." | Further illustrates self-attribution of harm, compounding reluctance to access professional support. |
| | *Ridicule and social absurdity* | *„[Die Menschen in meinem Umfeld denken] Davon kriegt man Psychosen und denkt man wäre eine Banane, irgendein Hippie, super peinlich.“* (Roman, Z. 108–123) | "[The people around me think] You get psychosis from it and think you're a banana — just some hippie, completely embarrassing." | Cultural framings oscillating between danger and ridicule were deeply internalised, producing shame that prevented open communication about symptoms. |
| | *Double stigma (drug + mental health)* | *„Aber bei psychischen Erkrankungen habe ich immer noch das Gefühl, das ist so stigmatisiert. Stell dich nicht so an oder so schlimm kann das doch nicht sein.“* (Lea, Z. 266–280) | "But with mental illness I still feel that it's so stigmatised. Don't be so dramatic, it can't be that bad." | Reflects multi-layered stigma combining drug-related moral judgement with broader societal devaluation of psychological suffering. |
| | *Self-blame — resigned* | *„Ja, selber Schuld, ne?“* (Tobias, Z. 128) | "Well, serves me right, doesn't it?" | A stark, condensed expression of internalised blame that forecloses the experience of suffering as a legitimate medical concern. |

| | | | | |
|---|---|---|---|---|
| **3.1 The Absence of Professional Understanding and Clinical Recognition** | *Lack of clinical knowledge* | *„Aber hat dann halt er gesagt, okay, das muss ich mal auf dem nächsten Kongress ansprechen. Also er wusste darüber dann nichts.“* (Selina, Z. 49–51) | "But then he said, okay, I'll have to bring this up at the next conference. So he simply had no knowledge of it at all." | Highlights epistemic gaps in clinical understanding of psychedelic-associated conditions, limiting diagnostic guidance and undermining therapeutic trust. |
| | *Moralising over clinical engagement* | *„Er [der Arzt] hatte ein bisschen wenig Zeit und kannte sich nicht so wirklich damit aus. [Er sagte:] 'Sowas sollte man nicht nehmen… Welche Pilze, was ist das denn?'“* (Marie, Z. 256–293) | "He [the doctor] had little time and didn't really know about it. [He said:] 'One shouldn't take things like that… Magic Mushrooms — what even is that?'" | Shows moralistic responses replacing clinical engagement, leaving the patient without either a diagnosis or a sense of being heard. |
| | *Anticipated incompetence* | *„Die sind halt auch alle nicht so auf so ein Zeug [Psychedelika] spezialisiert, habe ich da jetzt auch nicht so die Hoffnung gehabt.“* (Manuel, Z. 65–74) | "None of them are really specialised in that sort of thing [psychedelics] either, so I didn't have much hope there." | Anticipated clinical incompetence led to pre-emptive withdrawal from help-seeking, demonstrating how structural knowledge gaps erode trust before the clinical encounter. |
| | *Misdiagnosis — psychosis label* | *„[Mir wurde gesagt] Es ist eine Psychose. [Aber] Ich weiß ja, dass [beispielsweise] die Heizung normal fest ist [und sich nicht wirklich bewegt].“* (Julia, Z. 131–135) | "[I was told] It is a psychosis. [But] I know that [for example] the heater is normally fixed [and doesn't really move]." | Demonstrates premature psychosis labelling without phenomenological assessment, contradicting the patient's own preserved reality testing and intensifying loss of agency. |
| | *Epistemic injustice* | *„Googelt das mal selber… Ich wusste gar nicht, was [mit mir] los ist. Es wurde viel [mit Medikamenten] experimentiert. Ich wusste nicht, dass Menschen so leiden können.“* (Roman, Z. 133–173) | "Google it yourselves… I had no idea what was [happening to me]. There was a lot of [medication] experimentation. I didn't know that people could suffer so much." | Marks clinical dismissal and destabilising treatment practices. The patient's experiential knowledge was discounted while standard interventions worsened symptoms — a paradigmatic instance of epistemic injustice. |
| | *Compounded access barriers* | *„Du telefonierst dir den Arsch ab, du schreibst E-Mails und das halt in einem State, wo es dir so beschissen geht und jeder soziale Kontakt dir so viel Kraft raubt. So, und dazu halt dann auch noch mit dem Hintergedanken, wie offen kann ich da über generell Drogenkonsum halt sprechen.“* (Tobias, Z. 215–225) | "You spend hours on the phone, you write emails — and all that in a state where you feel so awful and every social contact drains so much energy. And on top of that with the background thought: how openly can I actually talk about drug use there." | Captures the compounded cost of navigating healthcare systems while severely symptomatic, intensified by anticipatory stigma regarding substance disclosure. |
| | *Systemic access barriers — waiting lists* | *„Ich stand bei 15 Therapeut:innen auf der Warteliste… [bis zum Therapieplatz hat es elf Monate gedauert].“* (Tobias, Z. 215–225) | "I was on the waiting list of 15 therapists… [it took eleven months until I got a therapy place]." | Illustrates systemic access barriers, likely compounded by stigma-related reluctance and the absence of specialised referral pathways for psychedelic-related conditions. |
| | *Loss of agency* | *„Keiner hört mir zu. Hilfe, Hilfe.“* (Julia, Z. 486–487) | "Nobody is listening to me. Help, help." | A condensed expression of epistemic invalidation that escalated into a profound sense of loss of agency within the healthcare system. |

| | | | | |
|---|---|---|---|---|
| | *Call for provider training* | *„Und es wäre schon irgendwie, glaube ich, ganz cool, wenn man da ein bisschen mehr Ahnung hätte, wie man das überhaupt behandelt."* (Marie, Z. 490–492) | "And I think it would be really great if there were a bit more knowledge out there about how to actually treat this." | Reflects participants' urgent calls for improved provider training and system-wide awareness of psychedelic-associated conditions. |
| **3.2 The Emergence of Self-Organised Support and Alternative Coping** | *Online communities as primary resource* | *„Die einzige wirkliche Anlaufstelle, worüber ich etwas erfahren konnte, war Reddit. Eine ganz große Community die recht aktiv ist und darüber postet, um darüber ein bisschen was zu verstehen."* (Max, Z. 53–55) | "The only real resource I could find anything out through was Reddit. A very large community that's quite active and posts about it — to try to understand it a little." | Shows the turn to peer-based, bottom-up knowledge repositories as the primary source of orientation when formal services fail. |
| | *Online orientation* | *„Das habe ich auf Reddit gelesen."* (Manuel, Z. 102–107) | "I read that on Reddit." | Reflects informational reliance on user-generated online communities in the absence of accessible clinical expertise. |
| | *Normalisation through peer comparison* | *„Und das, was ich dort gelesen habe, muss ich auch sagen, dann ist mein Fall wahrscheinlich auch nicht so schlimm."* (Marie, Z. 347–358) | "And reading what was posted there — I have to say, my case is probably not that bad." | Demonstrates relief through comparative peer narratives rather than clinical validation, illustrating the normalising and stabilising function of online support networks. |
| | *Supplement-based self-management* | *„Und dann habe ich gelesen, was kann man gegen Angst machen? Man kann Baldrian nehmen, dann GABA und das hat noch besser geholfen."* (Paul, Z. 263–270) | "And then I read about what you can do for anxiety. You can take valerian, then GABA and that helped even better." | Indicates pharmacological self-experimentation to regain a sense of control over symptoms in the absence of adequate medical guidance. |
| | *Lifestyle change* | *„Mit Sport, mit gesunder Ernährung angefangen."* (Max, Z. 74–75) | "Started with sports and eating more healthily." | Illustrates self-initiated holistic coping strategies as a substitute for formal treatment. |
| | *Mind-body practices* | *„Ganz viel Yoga gemacht, mit Sport angefangen."* (Luca, Z. 143–144) | "Did a lot of yoga, started with sports." | Embodies embodied coping approaches adopted when formal treatment options were exhausted or inaccessible. |
| | *Hermeneutic reframing* | *„Es kann gut sein, dass dieses LSD mich für diese Beziehungsdynamiken [mit meiner Ex-Partnerin] enorm empfänglich gemacht hat. Das wäre so meine Theorie dazu."* (Manuel, Z. 221–224) | "It's quite possible that the LSD made me enormously susceptible to those relationship dynamics [with my ex-partner]. That would be my theory about it." | Demonstrates construction of personal explanatory models to integrate psychedelic experiences into broader biographical contexts, regaining coherence in the absence of clinical frameworks. |
| **3.3 Helpful Clinical Support as a Restorative Resource** | *Enabling narrative openness* | *„Ja, okay, dann werde ich da ja schon frei von erzählen können, dass ich [Psychedelika] konsumiert habe."* (Tobias, Z. 205–206) | "Yes, okay, then I'll be able to talk freely about having used [psychedelics]." | Anticipated non-judgement facilitates disclosure of substance use, crucial for accurate clinical assessment. Reflects how empathic environments counteract self-stigma and enable therapeutic engagement. |

| | | | | |
|---|---|---|---|---|
| | *Diagnostic naming as relief* | *„Wo sie [die Psychiaterin] dann sofort erkannt hat, ah, okay, das ist mit dem Jungen los. Und dann habe ich selber auch gemerkt, aha, okay, das ist Derealisation nur und ich sterbe jetzt nicht, ich kriege keine Psychose und keine Ahnung was."* (Roman, Z. 36–41) | "Where she [the psychiatrist] immediately recognised, ah, okay, this is what's happening with the young man. And then I also realised, aha, okay, this is just derealisation and I'm not dying now, I'm not getting a psychosis or whatever." | Diagnostic naming provides cognitive containment. The clinician's rapid contextualisation transforms overwhelming phenomenological states into medically intelligible experiences, substantially reducing existential panic. |
| | *Reassurance and symptom reframing* | *„Und die haben mich dann auch noch mal beruhigt, dass ich mir keinen dauerhaften Schaden eingefangen habe deswegen. Und ich hab mir nichts mit dem LSD kaputt gemacht. Ich bin nicht verrückt geworden deswegen oder sonstiges."* (Manuel, Z. 68–73) | "And they also reassured me that I had not sustained any permanent damage because of it. And I hadn't broken anything with the LSD. I didn't go crazy because of it or anything like that." | Professional reassurance reduced fear and reframed symptoms, illustrating the clinically significant impact of expert validation on psychological stabilisation. |
| | *Feeling seen and understood* | *„Ja, [die Erfahrung mit meinem Psychiater war] auf jeden Fall positiv, weil er da halt sich gut auskennt. Und da hat man sich halt dann irgendwie so gesehen und verstanden gefühlt."* (Manuel, Z. 107–114) | "Yes, [the experience with my psychiatrist was] definitely positive, because he really knows his stuff there. And then you somehow felt seen and understood." | Professional familiarity with psychedelic phenomenology fosters a therapeutic alliance marked by recognition rather than judgement. Feeling 'seen' directly counteracts prior experiences of delegitimisation. |
| | *Non-dismissal of unusual perceptual content* | *„(Ich habe dem Psychiater erzählt) Ich habe das LSD genommen. Meine Stimme klingt mir fremd. Ich sehe hier Farben an der Wand. Und es wurde nicht abgeblockt, sondern… alles aufgenommen."* (Julia, Z. 559–563) | "(I told the psychiatrist) I took the LSD. My voice sounds strange to me. I see colours on the wall here. And it was not blocked, but… everything was taken in." | Openness to unusual perceptual content without immediate pathologising represents a restorative clinical stance, enabling disclosure of experiences typically met with dismissal. |
| | *Restored trust and continuity of care* | *„Aber irgendwann ging es halt gar nicht mehr. Dann war ich in der Klinik. Und dann habe ich eben gesagt, ja, komm, ich gebe dem Ganzen noch eine Chance und gehe dann zu dieser [Psychiaterin]."* (Roman, Z. 36–41) | "But at some point I simply couldn't go on anymore. Then I was admitted to the clinic. And then I said to myself, okay, come on, give it one more chance and go to this [psychiatrist]." | Following multiple negative encounters, a positive clinical relationship restored willingness to engage with formal care, underscoring the cumulative impact of prior dismissal on help-seeking motivation. |

**Column guide:** Theme = analytical theme from reflexive thematic analysis; Subtheme = lower-level analytic category; German Original = verbatim participant excerpt with pseudonym and transcript line number (Z.); English Translation = translation used in main text narrative (or provided here for supplementary-only excerpts); Interpretive Note = analytic commentary on the excerpt's significance within the theme.

**Abbreviations:** HPPD = Hallucinogen Persisting Perception Disorder (DSM-5: 292.89); DDD = Depersonalisation/Derealisation Disorder; LSD = lysergic acid diethylamide; Z. = Zeile (line number in verbatim transcript); [...] = researcher abridgement; [ ] = researcher clarification inserted into excerpt; all participant names are pseudonyms.
