## Supplemental: Coreq List for "Persisting Psychological Complications Following the Use of Classic Psychedelics: A Qualitative Study of Help-Seeking Experiences"

### Supplementary File: COREQ Checklist

*Consolidated Criteria for Reporting Qualitative Research (COREQ) – Tong A, Sainsbury P, Craig J (2007). Qualitative Health Research, 17(6), 1068–1082.*

#### Study: Persisting Psychological Complications Following the Use of Classic Psychedelics: A Qualitative Study of Help-Seeking Experiences

| Domain | No. | Topic | Guiding Question | Reported | Location in Manuscript |
|---|---|---|---|---|---|
| **Domain 1: Research Team and Reflexivity** | | | | | |
| Personal characteristics | | | | | |
| | 1 | **Interviewer / facilitator** | Which author/s conducted the interview or focus group? | YES | Methods §2.4: interviews conducted by (L.M.J); see also §2.7 Reflexivity |
| | 2 | **Credentials** | What were the researcher's credentials, e.g. PhD, MD? | YES | Author affiliations on title page; primary researcher owns a Master's Degree |
| | 3 | **Occupation** | What was their occupation at the time of the study? | YES | Methods §2.1: PPaPS project at Charité University Medicine Berlin |
| | 4 | **Gender** | Was the researcher's gender specified? | YES | Methods §2.7 (Reflexivity and Positionality). |
| | 5 | **Experience and training** | What experience or training did the researcher have? | YES | §2.7: prior professional/personal engagement with psychedelics noted; formal methodological training specified. |
| Relationship with participants | | | | | |
| | 6 | **Relationship established** | Was a relationship established prior to study commencement? | YES | §2.2: participants recruited from the PPaPS study database; no new recruitment relationship required. |
| | 7 | **Participant knowledge of the interviewer** | What did participants know about the researcher? e.g. personal goals, reasons for doing the research | YES | §2.4: interview context described; statement of what participants were told about the researcher's role is included. |
| | 8 | **Interviewer characteristics** | What characteristics were reported about the interviewer/facilitator? e.g. bias, assumptions, reasons for interest in the topic | YES | §2.7: detailed reflexivity/positionality statement; team's prior engagement with psychedelic research disclosed. |
| **Domain 2: Study Design** | | | | | |
| Theoretical framework | | | | | |
| | 9 | **Methodological orientation and theory** | What methodological orientation was stated to underpin the study? e.g. grounded theory, discourse analysis, ethnography, phenomenology, content analysis | YES | §2.1: Reflexive Thematic Analysis (Braun & Clarke, 2006, 2024); social-constructionist and hermeneutic epistemology stated. |
| Participant selection | | | | | |
| | 10 | **Sampling** | How were participants selected? e.g. purposive, convenience, consecutive, snowball | YES | §2.2: purposive sampling from PPaPS database; eligibility criteria stated. |
| | 11 | **Method of approach** | How were participants approached? e.g. face-to-face, telephone, mail, email | YES | §2.2: recruited from existing study database; contact method explicitly stated |

| | | | | | |
|---|---|---|---|---|---|
| | 12 | **Sample size** | How many participants were in the study? | YES | §2.2 & §3: n = 13 (of 25 eligible; response rate 52%). |
| | 13 | **Non-participation** | How many people refused to participate or dropped out? Reasons? | YES | §2.3: 12 of 25 eligible declined (response rate stated); reasons for non-participation not known. |
| Setting | | | | | |
| | 14 | **Setting of data collection** | Where was the data collected? e.g. home, clinic, workplace | YES | §2.4: encrypted video calls (ZOOM®). |
| | 15 | **Presence of non-participants** | Was anyone else present besides the participants and researchers? | YES | §2.4: individual interviews; no third parties mentioned. |
| | 16 | **Description of sample** | What are the important characteristics of the sample? e.g. demographic data, date | YES | §2 & Table 1: demographics (gender, age, education, diagnoses, interview duration, index experience). |
| Data collection | | | | | |
| | 17 | **Interview guide** | Were the questions, prompts, guides provided by the authors? Was it pilot tested? | YES | §2.4: four-block interview guide described; pilot testing not mentioned. |
| | 18 | **Repeat interviews** | Were repeat interviews carried out? If yes, how many? | YES | Single interviews conducted. No repeat interviews were intended. |
| | 19 | **Audio/visual recording** | Did the research use audio or visual recording to collect the data? | YES | §2.4: audio recorded; recordings deleted after transcription. |
| | 20 | **Field notes** | Were field notes made during and/or after the interview or focus group? | YES | §2.7: reflective memos documented during analysis. |
| | 21 | **Duration** | What was the duration of the interviews or focus group? | YES | §2.4 & Table 1: range 20–68 minutes; individual durations in Table 1. |
| | 22 | **Data saturation** | Was data saturation discussed? | YES | §2.3: thematic sufficiency discussed per Braun & Clarke (2019); no new conceptual themes in final 3 interviews. |
| | 23 | **Transcripts returned** | Were transcripts returned to participants for comment and/or correction? | YES | (§2.7); reason stated. |
| **Domain 3: Data Analysis and Reporting** | | | | | |
| Data analysis | | | | | |
| | 25 | **Description of the coding tree** | Did authors provide a description of the coding tree? | PARTIAL | §2.4: six-phase process described; no visual coding tree or codebook appended. Supplementary Table S1 partially fulfils this. |
| | 24 | **Number of data coders** | How many data coders coded the data? | YES | §2.4: coding discussed within PPaPS team; primary coder (L.M.J.) specified. |
| | 26 | **Derivation of themes** | Were themes identified in advance or derived from the data? | YES | §2.4: inductive coding; themes derived from data (bottom-up), contextualized with theory post-hoc. |
| | 27 | **Software** | What software, if applicable, was used to manage the data? | YES | Whisper.ai stated for transcription (§2.4); qualitative analysis software f4analyse (§2.5). |

| | | | | | |
|---|---|---|---|---|---|
| | **28** | **Participant checking** | Did participants provide feedback on the findings? | **YES** | §2.7: participants not involved in member-checking; reason explicitly stated and justified. |
| **Reporting** | | | | | |
| | **29** | **Quotations presented** | Were participant quotations presented to illustrate the themes/findings? | **YES** | Results (§3.1–3.3.3): extensive quotations with pseudonym and line number; German originals in Supplementary Table S1. |
| | **30** | **Data and findings consistent** | Was there consistency between the data presented and the findings? | **YES** | Themes supported by multiple quotations across participants; analytic interpretations align with excerpts presented. |
| | **31** | **Clarity of major themes** | Were major themes clearly presented in the findings? | **YES** | 3 themes with subthemes (§3.3 has 3 subthemes); each given a titled section with summary and illustrative quotes. |
| | **32** | **Clarity of minor themes** | Is there a description of diverse cases or discussion of minor themes? | **YES** | §3.1: recreational subgroup (≈31%) noted as a contrasting minority. |

**YES** = criterion fully met **PARTIAL** = criterion partially met / requires minor addition **NO** = criterion not met / information absent

*Reference: Tong A, Sainsbury P and Craig J (2007) Consolidated criteria for reporting qualitative research (COREQ): a 32-item checklist for interviews and focus groups. International Journal for Quality in Health Care 19(6): 349–357.*
